# Predominance of multidrug-resistant (MDR) bacteria causing urinary tract infections (UTIs) among symptomatic patients in East Africa: a call for action

**DOI:** 10.1101/2023.06.13.23291274

**Authors:** Antonio Maldonado-Barragán, Stephen E. Mshana, Katherine Keenan, Xuejia Ke, Stephen H. Gillespie, John Stelling, John Maina, Joel Bazira, Ivan Muhwezi, Martha F. Mushi, Dominique L. Green, Mike Kesby, Andy G. Lynch, Wilber Sabiiti, Derek J. Sloan, Alison Sandeman, John Kiiru, Benon Asiimwe, Matthew T. G. Holden, HATUA consortium

## Abstract

In low-and middle-income countries, antibiotics are often prescribed for patients with symptoms of urinary tract infections (UTIs) without microbiological confirmation. Inappropriate antibiotic use can contribute to antimicrobial resistance (AMR) and the selection of multi-drug resistant (MDR) bacteria. Data on antibiotic susceptibility patterns of cultured bacteria are important in drafting empirical treatment guidelines and monitoring resistance trends, which can prevent the spread of AMR. In East Africa, antibiotic susceptibility data are sparse. To fill the gap, this study reports common microorganisms and their susceptibility patterns isolated from patients with UTI-like symptoms in Kenya, Tanzania, and Uganda in 2019-2020. Microbiologically confirmed UTI was observed in 2,653 (35.0%) of the 7583 patients studied. The predominant bacteria were *Escherichia coli* (37.0%), *Staphylococcus* spp. (26.3%), *Klebsiella* spp. (5.8%) and *Enterococcus* spp. (5.5%). *E. coli* contributed 982 of the isolates with an MDR proportion of 52.2%. *Staphylococcus* spp. contributed 697 of the isolates with an MDR rate of 60.3%. The overall proportion of MDR bacteria (n=1,153) was 50.9%. MDR bacteria are common causes of UTI in patients attending healthcare centres in East African countries, which emphasizes the need for investment in laboratory culture capacities and diagnostic algorithms to improve accuracy of diagnosis that will lead to appropriate antibiotic uses to prevent and control AMR.

## 1. Introduction

Increase of antimicrobial resistance (AMR) is currently considered one of the top 10 global public health threats [1]. In 2019, there were an estimated 4.95 million deaths associated with antibacterial resistance (ABR) including 1.27 million deaths directly attributable to ABR [2]. Among all world regions, Sub-Saharan Africa has the largest burden of ABR-attributable deaths [2], although most contemporary ABR estimates in that region are based on incredibly sparse data [2,3,4]. This serious threat requires a better assessment of ABR to understand the current and future burden of AMR and to direct the use of antibiotics (ABs) more effectively. This motivated the formation of the interdisciplinary consortium “Holistic Approach to Unravel Antibacterial Resistance in East Africa” (HATUA), which aimed to explore the burden and drivers of ABR associated with urinary tract infections (UTIs) in three East African countries, Kenya, Tanzania, and Uganda [5].

UTI is an inflammatory response of the urothelium to bacterial invasion and is considered the most frequent community-acquired bacterial infection in the world, affecting more than 150 million people per year [6,7]. In addition, UTIs are the third most frequent healthcare-associated infections (HAIs), with approximately one-third of all deaths associated to HAIs [8]. Globally, deaths attributable to and associated with ABR in UTIs in 2019 were approximately 65,000 and 250,000, respectively [2,7]. UTI is the second most frequent reason for using ABs in the community, which can contribute to the emergence of multi-drug resistant (MDR) bacteria [9]. The prevalence of MDR bacteria – defined as bacteria with nonsusceptibility to at least one agent in ≥ 3 antimicrobial categories [10] – associated with UTI has increased worldwide, thus limiting the therapeutic options for the treatment of infections caused by those microorganisms [11,12,13].

In community-acquired UTIs, AB treatment is usually prescribed empirically. The selection of the empirical AB is based on surveillance mechanisms addressing the frequency of uropathogens and their antimicrobial resistance profiles. However, culture and susceptibility data for community UTI infections are unavailable in many LIMCs regions such as East Africa, mainly due to limited health service funding, and paucity of microbiology laboratory capacities including limited skilled personnel [2,3,4]. These data are critical for prescribing the appropriate empirical AB, which could contribute to reducing the emergence of MDR bacteria and therefore UTI-associated complications such as pyelonephritis or bacteraemia through more effective treatment [15].

The main goals of this study are, therefore, to describe the proportion of microbiologically confirmed UTI in symptomatic patients who attended clinics in Kenya, Tanzania, and Uganda, to characterize the main uropathogenic bacteria responsible and their antimicrobial resistance profiles, and to estimate the proportion of MDR bacteria associated with UTIs. The findings presented here can input to UTI empirical treatment guidelines in East Africa, helping to prevent the AMR-associated complications and deaths.

## 2. Methods

### 2.1. Study design, patient selection, and sample size

The sample collection took place between April 2019 and November 2020 in Kenya, Tanzania, and Uganda in different levels of health facilities and locations (Table S1). The study included adults and children (≥ 2 years) with signs and symptoms of UTI (detailed description for inclusion of patients are shown in the supplementary material (Method S1). Self-collected mid-stream clean catch urine samples were obtained from each patient, as described previously [5]. Patients were classified according to their stay at the recruitment health facilities as outpatient (visits with no overnight stay) or inpatient (overnight or longer stay). A total of 7,583 patients with symptomatic UTI were recruited from Kenya (n=1,903), Tanzania (n=3,852), and Uganda (n=1,828) (Figure 1).

**Figure 1.**
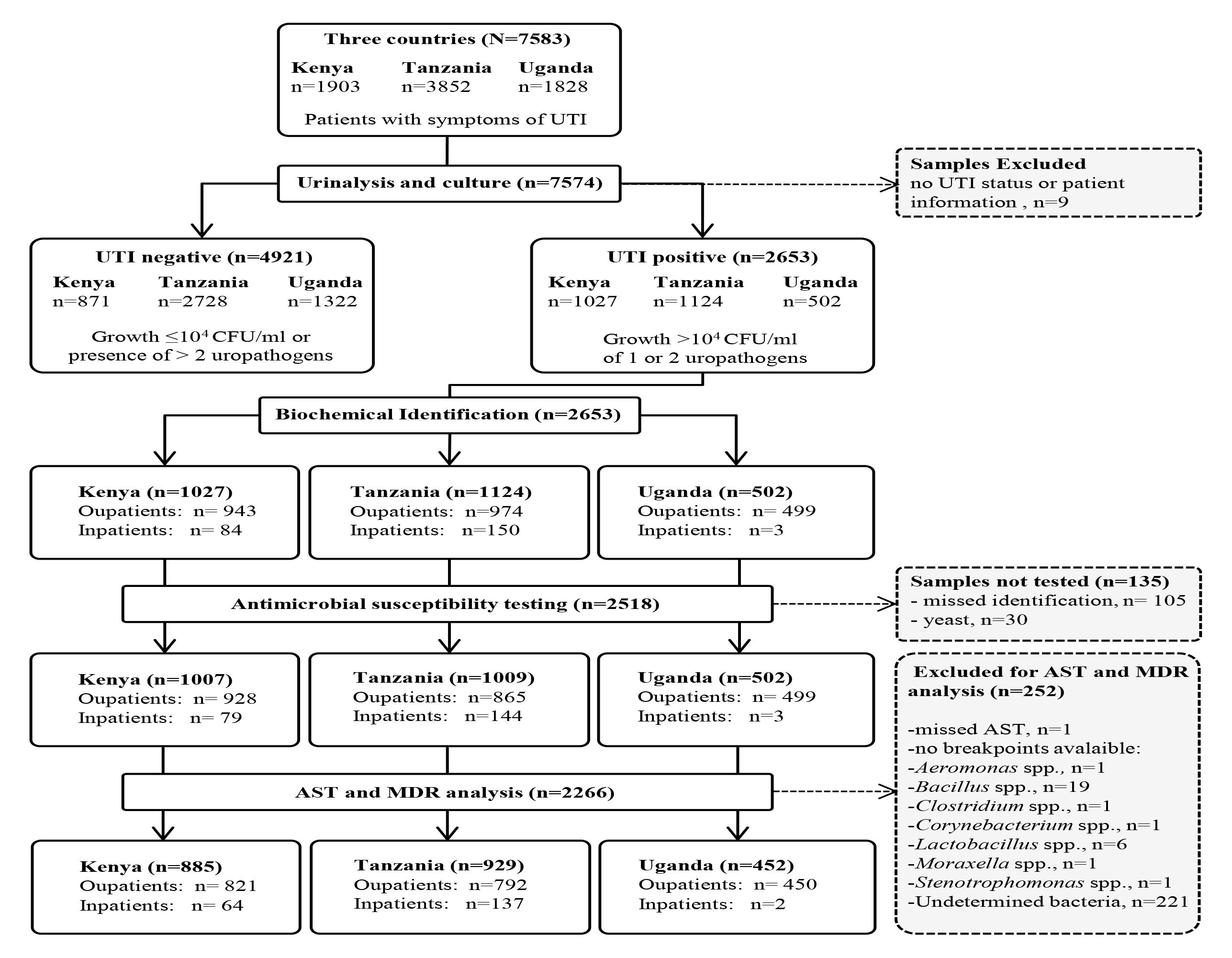
Consort diagram describing HATUA patient recruitment and processing and analysis of their urine samples.

### 2.2. Urine culture and biochemical identification of isolates

A standard disposable sterile plastic loop was used to inoculate 1μl or 10μl of mid-stream urine sample onto cysteine–lactose–electrolyte–deficient (CLED), sheep blood (SBA), and MacConkey agar plates (Oxoid, Basingstoke, UK) [16]. After 18-24 hours of incubation at 37°C under aerobic conditions, cultures were quantified. Microbiologically confirmed urinary tract infection (hereafter UTI positive samples) was defined by the presence of >10^4^ colony-forming units per millilitre of one or two uropathogens. Contaminated samples (> 10^4^ CFU/mL growth of more than two different uropathogens or any growth of < 10^4^ CFU/mL) and those with no microbial growth were considered UTI negative. In samples containing two possible uropathogens, only the predominant or the most probable uropathogen (subjected to the evaluation by an experienced clinical microbiologist) was included in the analysis of the data.

In-house methods were used to identify Gram-negative bacteria and included: colonial morphology on CLED, Blood, and MacConkey agar (Oxoid, Basingstoke, UK), and Triple Sugar Iron agar, Sulphur Indole and Motility, Citrate, Oxidase, Urease, Voges-Proskauer and Methyl Red tests. Coagulase, Catalase, Bile-esculin, and Bacitracin-Sulfamethoxazole disc susceptibility tests were used to confirm the presence of Gram-positive bacteria, which were identified using colony morphology on SBA.

### 2.3. Antimicrobial susceptibility testing

Antimicrobial susceptibility testing (AST) was performed by the conventional Kirby–Bauer disk diffusion method according to Clinical and Laboratory Standards Institute (CLSI) M02 document [17]. The discs (Oxoid, Basingstoke, UK) tested were ampicillin (10µg), amoxicillin/clavulanic acid (20/10µg), cefoxitin (30µg), tetracycline (30µg), trimethoprim (5µg), ciprofloxacin (5µg), gentamicin (10µg), nitrofurantoin (100µg), ceftriaxone (30µg), ceftazidime (30µg), erythromycin (15µg), linezolid (10µg) and vancomycin (30µg). The susceptibility or non-susceptibility (resistance) to the tested ABs was determined by using the breakpoints (zone diameter interpretive criteria) indicated in the M100 document of CLSI guidelines [18], as further detailed in supplementary Method S2. Those isolates that showed intermediate resistance to a given AB were considered resistant to such AB. Prediction of possible extended-spectrum β-lactamase (ESBL) producers was based on ceftazidime and/or ceftriaxone resistance, following the criteria indicated in the CLSI guidelines [18].

### 2.4. Definition and analysis of multidrug resistance

MDR bacteria were defined as isolates resistant to at least one agent in three or more classes of antimicrobial agents, following the European Centre for Disease Prevention and Control (ECDC) guidelines [10], with some modifications as specified in supplementary Method S3. MDR rates were calculated by considering the number of MDR isolates divided by the total isolates where valid MDR data were obtained.

### 2.5. Patient characteristics

A questionnaire was conducted with all patients (or their parents/guardians) which captured sociodemographic factors including age, gender, and other factors (e.g. education, marital status and household socioeconomic factors). Selected variables are shown in Table 1.

**Table 1.** Characteristics of the patients with symptoms of UTI at the time of recruitment

| **Variables/Country** | **Kenya**<br>**n (%)** | **Tanzania**<br>**n (%)** | **Uganda**<br>**n (%)** | **Total**<br>**n (%)** |
| --- | --- | --- | --- | --- |
| **Patient type** |  |  |  |  |
| Outpatient | 1754(92.4) | 3552(92.2) | 1815(99.5) | 7121(94.0) |
| Inpatient | 144(7.6) | 300(7.8) | 9(0.5) | 453(6.0) |
| **Gender** |  |  |  |  |
| Male | 348(18.3) | 1097(28.5) | 295(16.2) | 1740(23.0) |
| Female | 1550(81.7) | 2754(71.5) | 1529(83.8) | 5833(77.0) |
| Missing | NA | 1(0.0) | NA | 1(0.0) |
| **Age in years** |  |  |  |  |
| <18 | 79(4.2) | 343(8.9) | 62(3.4) | 484(6.4) |
| 18-24 | 493(26.0) | 718(18.6) | 573(31.4) | 1784(23.6) |
| 25-34 | 837(44.1) | 950(24.7) | 573(31.4) | 2360(31.2) |
| 35-44 | 294(15.5) | 550(14.3) | 304(16.7) | 1148(15.2) |
| 45-54 | 97(5.1) | 425(11.0) | 173(9.5) | 695(9.2) |
| 55-64 | 43(2.3) | 321(8.3) | 71(3.9) | 435(5.7) |
| 65-74 | 35(1.8) | 281(7.3) | 43(2.4) | 359(4.7) |
| 75 and above | 20(1.1) | 261(6.8) | 22(1.2) | 303(4.0) |
| Missing | 0(0) | 3(0.1) | 3(0.2) | 6(0.1) |
| **Hospital level<sup>1</sup>** |  |  |  |  |
| Level 2 (low) | 0(0) | 309 (8.0) | 394(21.6) | 703(9.3) |
| Level 3 | 384(20.2) | 2152(55.9) | 1023(56.1) | 3559(47.0) |
| Level 4 | 486(25.6) | 366(9.5) | 143(7.8) | 995(13.1) |
| Level 5 /6 (high) | 1028(54.2) | 1025(26.6) | 263(14.4) | 2316(30.6) |
| Missing | 0(0) | 0(0) | 1(0.1) | 1(0.0) |
| **TOTAL** | 1898 (100.0) | 3852 (100.0) | 1824 (100.0) | 7574 (100.0) |

<sup>1</sup>In all three countries, lower levels (1-3) refer to primary care, dispensaries, or community health centres. Level 4 typically refers to primary referral facilities or specialist health care facilities. Level 5 (and 6 in Kenyan) are higher level/tertiary facilities.

### 2.6. Data management and analysis

Data were captured using paper forms and electronically, using Epicollect5 mobile application (https://five.epicollect.net) [19]. Urinalysis, AST and MDR data was linked to the questionnaire data using anonymous patient identifiers. AB susceptibility/MDR rates were calculated in R Statistical Software (v4.1.1; R Core Team 2021). Descriptive analysis and χ2-testing with false discovery rate correction (Benjamini and Yekutieli 2001) were conducted in STATA 16 (StataCorp. 2019. Stata Statistical Software: Release 16. College Station, TX: StataCorp LLC).

### 2.7. Quality control

*Escherichia coli* ATCC 25922, *E. coli* NCTC 13353 (CTX-M-15 ESBL producer), *Staphylococcus aureus* ATCC 25923, *S. aureus* NCTC 13552 (*mecC*; MRSA), *Pseudomonas aeruginosa* ATCC 49189, *Proteus mirabilis* NCTC 10975, and *Enterococcus faecium* ATCC 51559 (*vanA*; vancomycin resistant) were used as reference strains for quality control of culture, biochemical identification, and antimicrobial susceptibility tests.

## 3. Results

### 3.1. Study participants and samples

A consort diagram of patient recruitment and analysis is shown in Figure 1. A total of 7,583 urine samples from non-repetitive patients with suspected UTI were collected in Kenya, Tanzania, and Uganda, of which 7,574 were categorized as either UTI-negative or UTI-positive, according to the results of the urine cultures. Of a total of 2,653 biochemically identified isolates, we obtained a valid AST result for 2,357 bacteria, which were subsequently included in the AST and MDR analysis.

### 3.2. Demographic features

Participant characteristics are shown in Table 1. Most were adult outpatients (89.9%) and females (77%). The modal age category was 25 to 34 years, and those aged 18-34 years contributed more than half (54.7%) of the total sample.

### 3.3. Proportion of microbiologically confirmed UTI

The overall proportion of microbiologically confirmed UTI across the three countries was 35.0%, being significantly higher in inpatients than in outpatients, in females, in patients recruited in higher-level facilities, and among patients over 65 years old (Table S3). Kenya reported a UTI proportion of 54.1%, which was higher than the proportion of 29.2% and 27.5% found in Tanzania and Uganda, respectively (Table S3).

### 3.4. Identity of isolates from UTI

A total of 2,653 isolates were characterised from urine samples of UTI-positive patients, 2,416 from outpatients and 237 from inpatients, of which 94.9% corresponded to bacteria, 1.1% to yeast, and 4.0% to isolates whose biochemical identification was not available. Among the bacterial isolates (n=2,518), 62.7% and 37.3% were Gram-negative and Gram-positive bacteria, respectively, of which 91.2% (n=2,297) were identified to at least the genus level.

Considering the three countries together (Table 2), *E. coli* was the predominant species (37.0%), followed by *Staphylococcus* spp. (26.3%), *Klebsiella* spp. (5.8%), and *Enterococcus* spp. (5.5%). By country, Kenya showed higher proportion of *Staphylococcus* spp. than Tanzania and Uganda, while Uganda showed higher proportion of *E. coli* than Kenya and Tanzania. Globally, *E. coli*, *Staphylococcus* spp., *Enterococcus* spp. and *Pseudomonas* spp. were more represented in samples from outpatients than inpatients, while proportions of *Klebsiella* spp. and *Acinetobacter* spp. were higher in samples from inpatients (Table S4).

**Table 2.** Distribution of significant microorganisms isolated from specimens of symptomatic patients with UTI (UTI positive patients), according to the country.

| Country | Kenya |  |  | Tanzania |  |  | Uganda |  |  | All 3 countries |  |  |
| --- | --- | --- | --- | --- | --- | --- | --- | --- | --- | --- | --- | --- |
| Microbial Isolates | n | %<sup>1</sup> | Prev (%)<sup>2</sup> | n | % | Prev (%) | n | % | Prev (%) | n | % | Prev (%) |
| *Escherichia coli* | 317 | 30.9 | 16.7 | 402 | 35.8 | 10.4 | 263 | 52.4 | 14.4 | 982 | 37.0 | 13.0 |
| *Klebsiella* spp. | 0 | 0 | 0 | 90 | 8.0 | 2.3 | 64 | 12.7 | 3.5 | 154 | 5.8 | 2.0 |
| *Proteus* spp. | 69 | 6.7 | 3.6 | 13 | 1.2 | 0.3 | 15 | 3.0 | 0.8 | 97 | 3.7 | 1.3 |
| *Acinetobacter* spp. | 8 | 0.8 | 0.4 | 19 | 1.7 | 0.5 | 5 | 1.0 | 0.3 | 32 | 1.2 | 0.4 |
| *Pseudomonas* spp. | 8 | 0.8 | 0.4 | 41 | 3.6 | 1.1 | 1 | 0.2 | 0.1 | 50 | 1.9 | 0.7 |
| Miscellaneous Gram-negative<sup>3</sup> | 101 | 9.8 | 5.3 | 111 | 9.9 | 2.9 | 52 | 10.4 | 2.9 | 264 | 10.0 | 3.5 |
| *Staphylococcus* spp. | 387 | 37.7 | 20.4 | 220 | 19.6 | 5.7 | 91 | 18.1 | 5.0 | 698 | 26.3 | 9.2 |
| *Enterococcus* spp. | 86 | 8.4 | 4.5 | 58 | 5.2 | 1.5 | 3 | 0.6 | 0.2 | 147 | 5.5 | 1.9 |
| Miscellaneous Gram-positive<sup>4</sup> | 31 | 3.0 | 1.6 | 55 | 4.9 | 1.4 | 8 | 1.6 | 0.4 | 94 | 3.5 | 1.2 |
| Yeast | 2 | 0.2 | 0.1 | 28 | 1.1 | 0.7 | 0 | 0.0 | 0.0 | 30 | 1.1 | 0.4 |
| Missing species data | 18 | 1.8 | 0.9 | 87 | 7.7 | 2.3 | 0 | 0.0 | 0.0 | 105 | 4.0 | 1.4 |
| Total | 1027 | 100 | 54.1 | 1124 | 100 | 29.2 | 502 | 100 | 27.5 | 2653 | 100 | 35.0 |

<sup>1</sup>% = percentage of isolates corresponding to that species, from that country (for example in the first column, calculated by 317/1027\*100).
<sup>2</sup>Prev = prevalence proportion (e.g. number of *E. coli* isolates with respect to the total number of urine specimens that were cultured in that country). For example, the third column is calculated by 317/1898\*100.
<sup>3</sup> Across all three countries, this comprises *Aeromonas* spp. (n=1), *Citrobacter* spp. (n=16), *Enterobacter* spp. (n=24), *Moraxella* spp. (n=1), *Morganella* spp. (n=6), *Pantoea* spp. (n=2), *Providencia* spp. (n=2), *Salmonella* spp. (n=2), *Serratia* spp. (n=4), *Shigella* spp. (n=1), *Stenotrophomonas* spp. (n=1), and undetermined Gram-negative bacteria (n=113)
<sup>4</sup> Across all three countries, this comprises *Bacillus* spp. (n=19), *Clostridium* spp. (n=1), *Corynebacterium* spp. (n=1), *Lactobacillus* spp. (n=6), *Streptococcus* spp. (n=50), and undetermined Gram-positive bacteria (n=17).

### 3.5. Regional burden of MDR in UTI pathogens

Of a total of 2,266 isolates included in the AST and MDR analysis (Figure 1), n=1,153 (50.9%) were categorized as MDR. By country, MDR rates were similar in Tanzania (60.9%) and Uganda (57.5%) while Kenya had a lower MDR rate (36.9%) (Table 3). Considering all countries together, the proportion of uropathogens that were classified as MDR was significantly higher in isolates from inpatients, those recruited in lower-level facilities, and in male patients (Table 3). By country, MDR proportions in Kenya and Tanzania were higher in males than in females, but this relationship was reversed in Uganda. By pathogen, *Staphylococcus* spp. showed the higher rates of MDR (60.3%), followed by *E. coli* (52.2%), *Klebsiella* spp. (50.6%), *Enterococcus* spp. (38.1%), and other *Enterobacterales* (31.2%) (Table 4). Within each pathogen group, isolates from inpatients or males exhibited higher MDR rates than isolates from outpatients and females, respectively (Table 4).

**Table 3.**
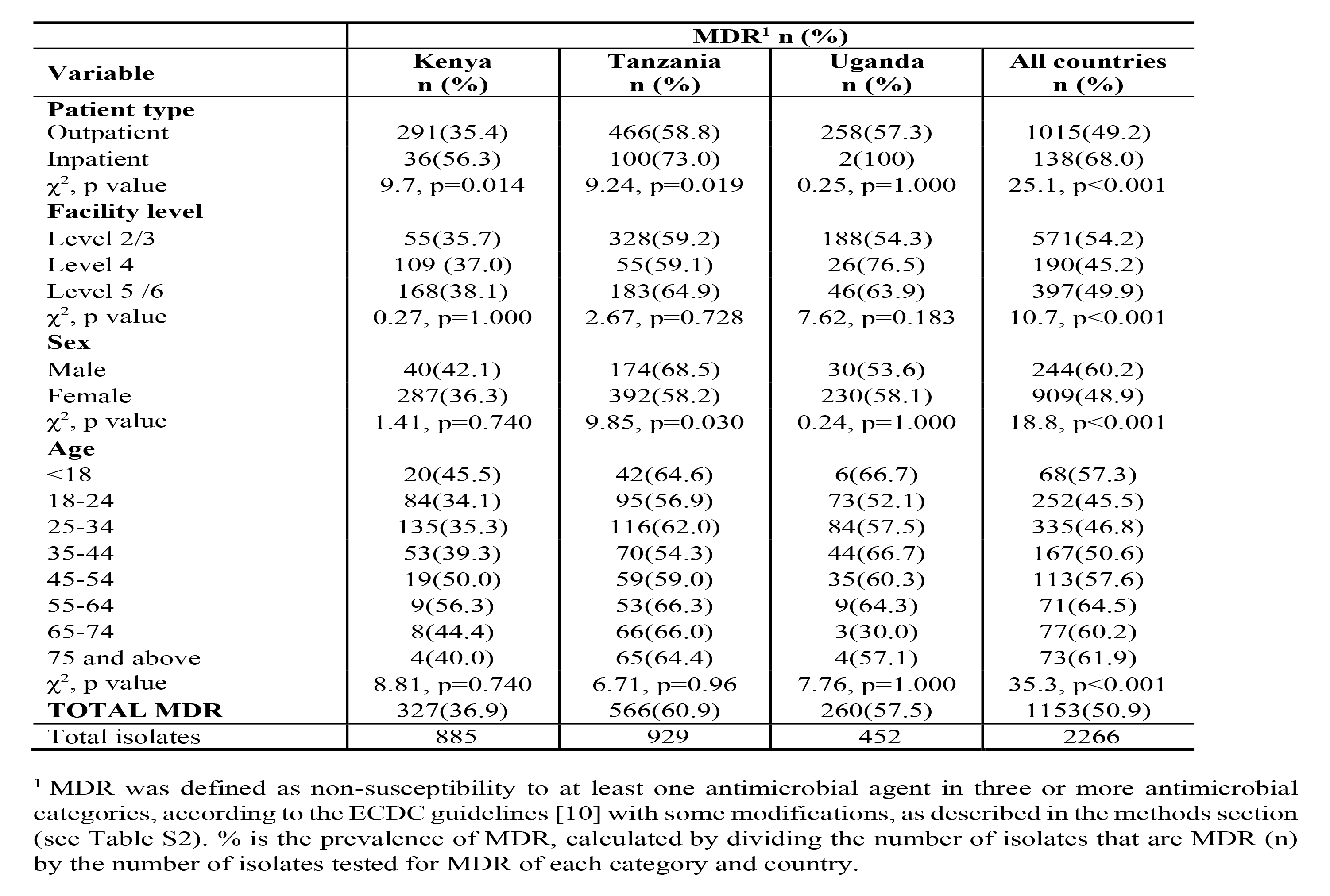
Prevalence of multi-drug resistant (MDR) bacteria in UTI positive samples by country, and according to patient type, hospital level, gender, and age.

| Variable | MDR<sup>1</sup> n (%) |  |  |  |
| --- | --- | --- | --- | --- |
|  | Kenya<br>n (%) | Tanzania<br>n (%) | Uganda<br>n (%) | All countries<br>n (%) |
| **Patient type** |  |  |  |  |
| Outpatient | 291(35.4) | 466(58.8) | 258(57.3) | 1015(49.2) |
| Inpatient | 36(56.3) | 100(73.0) | 2(100) | 138(68.0) |
| $\chi^2$, p value | 9.7, p=0.014 | 9.24, p=0.019 | 0.25, p=1.000 | 25.1, p<0.001 |
| **Facility level** |  |  |  |  |
| Level 2/3 | 55(35.7) | 328(59.2) | 188(54.3) | 571(54.2) |
| Level 4 | 109 (37.0) | 55(59.1) | 26(76.5) | 190(45.2) |
| Level 5 /6 | 168(38.1) | 183(64.9) | 46(63.9) | 397(49.9) |
| $\chi^2$, p value | 0.27, p=1.000 | 2.67, p=0.728 | 7.62, p=0.183 | 10.7, p<0.001 |
| **Sex** |  |  |  |  |
| Male | 40(42.1) | 174(68.5) | 30(53.6) | 244(60.2) |
| Female | 287(36.3) | 392(58.2) | 230(58.1) | 909(48.9) |
| $\chi^2$, p value | 1.41, p=0.740 | 9.85, p=0.030 | 0.24, p=1.000 | 18.8, p<0.001 |
| **Age** |  |  |  |  |
| <18 | 20(45.5) | 42(64.6) | 6(66.7) | 68(57.3) |
| 18-24 | 84(34.1) | 95(56.9) | 73(52.1) | 252(45.5) |
| 25-34 | 135(35.3) | 116(62.0) | 84(57.5) | 335(46.8) |
| 35-44 | 53(39.3) | 70(54.3) | 44(66.7) | 167(50.6) |
| 45-54 | 19(50.0) | 59(59.0) | 35(60.3) | 113(57.6) |
| 55-64 | 9(56.3) | 53(66.3) | 9(64.3) | 71(64.5) |
| 65-74 | 8(44.4) | 66(66.0) | 3(30.0) | 77(60.2) |
| 75 and above | 4(40.0) | 65(64.4) | 4(57.1) | 73(61.9) |
| $\chi^2$, p value | 8.81, p=0.740 | 6.71, p=0.96 | 7.76, p=1.000 | 35.3, p<0.001 |
| **TOTAL MDR** | 327(36.9) | 566(60.9) | 260(57.5) | 1153(50.9) |
| Total isolates | 885 | 929 | 452 | 2266 |

<sup>1</sup> MDR was defined as non-susceptibility to at least one antimicrobial agent in three or more antimicrobial categories, according to the ECDC guidelines [10] with some modifications, as described in the methods section (see Table S2). % is the prevalence of MDR, calculated by dividing the number of isolates that are MDR (n) by the number of isolates tested for MDR of each category and country.

**Table 4.** Prevalence of multi-drug resistant (MDR) bacteria in UTI positive samples for selected species, according to patient type, age, and gender.

|  | MDR<sup>1</sup> n (%) |  |  |  |  |
| --- | --- | --- | --- | --- | --- |
|  | *Escherichia coli* | *Klebsiella spp.* | Other<br>*Entero-*<br>*bacteriales*<sup>2</sup> | *Staphylococcus*<br>*spp.* | *Enterococcus*<br>*spp.* |
| **Patient type** |  |  |  |  |  |
| Outpatient | 460 (50.8) | 57 (44.9) | 39 (28.3) | 389 (59.5) | 45 (34.4) |
| Inpatient | 53 (69.7) | 21 (77.8) | 9 (56.3) | 31 (72.1) | 11 (68.8) |
| **$\chi^2$ p value** | 9.36, p=0.006 | 8.36, p=0.020 | 4.01, p=0.120 | 2.11, p=0.803 | 5.58, p=0.064 |
| **Age** |  |  |  |  |  |
| Adult | 481 (51.7) | 71 (49.7) | 43 (30.9) | 403 (59.9) | 52 (37.7) |
| Child | 31 (62.0) | 6 (60.0) | 5 (33.3) | 17 (70.8) | 4 (44.4) |
| **$\chi^2$ p value** | 1.64, p=0.367 | 0.09, p=1.000 | 0.01, p=1.000 | 0.71, p=0.970 | 0.01, p=1.000 |
| **Gender** |  |  |  |  |  |
| Male | 107 (71.3) | 37 (63.8) | 23 (44.2) | 43 (64.2) | 17 (58.6) |
| Female | 406 (48.8) | 41 (42.7) | 25 (24.5) | 377 (59.8) | 39 (33.1) |
| **$\chi^2$ p value** | 25.7, p<0.001 | 5.61, p=0.048 | 5.35, p=0.113 | 0.39, p=0.970 | 5.14, p=0.064 |
| **Total MDR** | 513 (52.2) | 78 (50.6) | 48 (31.2) | 420 (60.3) | 56 (38.1) |
| Total isolates | 982 | 154 | 154 | 697 | 147 |

<sup>1</sup> MDR was defined as non-susceptibility to at least one antimicrobial agent in three or more antimicrobial categories, according to the ECDC guidelines [10] with some modifications, as described in the methods section (see Table S2). % is the prevalence of MDR, calculated by dividing the number of isolates that are MDR (n) by the number of isolates tested for MDR of each category and selected species.
<sup>2</sup>Other *Enterobacterales* includes *Citrobacter*, *Enterobacter*, *Morganella*, *Proteus*, *Providencia*, *Pantoea*, *Salmonella*, *Serratia* and *Shigella* species.

### 3.6. Antibiotic susceptibility and MDR in *Enterobacterales*

The overall resistance rates of *Enterobacterales* ranged from 71.6% for trimethoprim to 7.5% for nitrofurantoin. The proportion of isolates with an ESBL and MDR were 31.4% and 49.5%, respectively (Table 5). Within bacterial groups, the resistance rates of the *E. coli* isolates ranged from 74.4% for trimethoprim to 4.1 % for nitrofurantoin (Table 5), with an ESBL and MDR proportion of 29.3% and 52.2%, respectively. *Klebsiella* spp. isolates exhibited resistance rates between 93.5% for ampicillin to 14.3% for nitrofurantoin (Table 5) and ESBL and MDR rates of 53.9% and 50.6%, respectively. The resistance rates of other *Enterobacterales* ranged from 61.8% for trimethoprim to 15.1% for gentamicin, displaying ESBL and MDR rates of 21.7% and 30.9%, respectively.

**Table 5.** Antibiotic susceptibility, ESBL and MDR rates of *Enterobacterales* and relevant Gram-positive uropathogens

|  | *Enterobacterales* |  | *Escherichia coli* |  | *Klebsiella* spp. |  | Other *Enterobacterales*<sup>1</sup> |  | *Staphylococcus* spp. |  | *Enterococcus* spp. |  |
| --- | --- | --- | --- | --- | --- | --- | --- | --- | --- | --- | --- | --- |
|  | %<sup>2</sup> | n<sup>3</sup> | % | n | % | n | % | n | % | n | % | n |
| **Ampicillin** | 65.9 | 1289 | 64.5 | 981 | 93.5 | 154 | 46.8 | 154 | NA<sup>4</sup> | NA | NA | NA |
| **Amoxicillin/**<br>**clavulanic acid** | 40.7 | 1276 | 38.1 | 970 | 60.1 | 153 | 37.3 | 153 | NA | NA | NA | NA |
| **Ceftazidime** | 24.0 | 1277 | 22.7 | 971 | 41.2 | 153 | 15.7 | 153 | NA | NA | NA | NA |
| **Ceftriaxone** | 30.2 | 1287 | 28.5 | 979 | 51.3 | 154 | 20.1 | 154 | NA | NA | NA | NA |
| **Ciprofloxacin** | 44.8 | 1289 | 45.8 | 982 | 43.5 | 154 | 39.2 | 153 | 38.2 | 696 | 40.1 | 147 |
| **Gentamicin** | 21.5 | 1170 | 22.1 | 900 | 24.8 | 129 | 14.9 | 141 | 20.9 | 669 | NA | NA |
| **Nitrofurantoin** | 7.5 | 1286 | 4.1 | 978 | 14.3 | 154 | 22.7 | 154 | 5.5 | 692 | 9.5 | 147 |
| **Trimethoprim** | 71.6 | 1289 | 74.4 | 981 | 63.0 | 154 | 62.3 | 154 | 81.8 | 696 | NA | NA |
| **Cefoxitin** | NA | NA | NA | NA | NA | NA | NA | NA | 39.7 | 692 | NA | NA |
| **Linezolid** | NA | NA | NA | NA | NA | NA | NA | NA | 15.9 | 661 | 8.8 | 136 |
| **Erythromycin** | NA | NA | NA | NA | NA | NA | NA | NA | 72.5 | 662 | 69.8 | 139 |
| **Tetracycline** | NA | NA | NA | NA | NA | NA | NA | NA | 48.0 | 671 | 50.3 | 145 |
| **Vancomycin** | NA | NA | NA | NA | NA | NA | NA | NA | NA | NA | 37.2 | 137 |
| **ESBL<sup>5</sup>** | 31.4 | 1290 | 29.3 | 982 | 53.9 | 154 | 22.1 | 154 | NA | NA | NA | NA |
| **MDR<sup>6</sup>** | 49.5 | 1290 | 52.2 | 982 | 50.6 | 154 | 31.2 | 154 | 60.3 | 697 | 38.1 | 147 |

<sup>1</sup> Other *Enterobacterales* includes *Citrobacter*, *Enterobacter*, *Morganella*, *Proteus*, *Providencia*, *Pantoea*, *Salmonella*, *Serratia* and *Shigella* species.
<sup>2</sup> % is the frequency of non-susceptible isolates, expressed in percentage.
<sup>3</sup> n is the total number of isolates that were tested for a specific antibiotic.
<sup>4</sup> not applicable (NA), the antibiotic was not tested in those isolates.
<sup>5</sup> Possible producers of extended-spectrum β-lactamase (ESBL) were determined by considering the resistance to antibiotics Ceftazidime and Ceftriaxone, according to CLSI guidelines [17].
<sup>6</sup> Multi drug-resistance (MDR) was defined as non-susceptibility to at least one antimicrobial agent in three or more antimicrobial categories, according to the ECDC guidelines [10] with some modifications, as described in the methods section.

*E. coli* from Kenya were less likely to be resistant to ampicillin, amoxicillin/clavulanic acid, trimethoprim, ciprofloxacin, ceftriaxone, and ceftazidime than those from Tanzania and Uganda, while in Tanzania, *E. coli* resistance to nitrofurantoin was higher than the other countries (Table S5). In addition, MDR and ESBL were less common among *E. coli* isolates from Kenya than those from Tanzania and Uganda, while MDR *Klebsiella spp.* were less represented in Uganda than in Tanzania. Regarding other *Enterobacterales*, isolates from Kenya were significantly less likely to be resistant to ampicillin, amoxicillin/clavulanic acid, ceftriaxone and ceftazidime than those from Tanzania and Uganda, and also showed lower ESBL and MDR rates. Ugandan isolates showed significantly higher rates of resistance to nitrofurantoin than isolates from other countries (Table S5).

The proportion of resistant isolates was generally higher in inpatients (Table S6) than outpatients (Table S7). Prevalence of ESBL and MDR among inpatient isolates was higher among *E. coli*, *Klebsiella* spp. and other *Enterobacterales* than those from outpatients (Table S6 and S7).

### 3.7. Antibiotic susceptibility and MDR in Staphylococci and Enterococci

The proportion of resistant *Staphylococcus* spp. isolates ranged from 5.5 % for nitrofurantoin to 81.8% for trimethoprim, with a MDR prevalence of 60.3% (Table 5). The cefoxitin resistance indicating methicillin resistance among staphylococci was 37.5%, 42.4% and 42.9% for Kenya, Tanzania, and Uganda respectively (Table S8). *Staphylococcus* spp. from Kenya showed a higher proportion of linezolid-resistant isolates (23.4%) than the other two countries (5.5%-7.5%) (Table S8). Isolates from Tanzania had the greatest proportion with MDR (72.3%).

For *Enterococcus* spp., the overall resistance rates ranged from 8.8% for linezolid to 69.8% for erythromycin, with a MDR prevalence of 38.1% (Table 5). Comparisons among countries revealed that *Enterococcus* sp. isolates from Kenya were less resistant to tetracycline and nitrofurantoin, and more resistant to linezolid than isolates from Tanzania and Uganda, with no significant differences in MDR rates (Table S8).

*Staphylococcus* spp. isolates from inpatients (Table S9) showed higher resistance than isolates from outpatients (Table S10), except for ABs ciprofloxacin, trimethoprim and tetracycline, displaying also increased MDR (72.1% vs. 59.5%) (Tables S9 and S10). *Enterococcus* spp. from inpatients were more resistant to ciprofloxacin, erythromycin and tetracycline, and showed higher MDR than outpatients (68.8% vs. 34.4%) (Tables S9 and S10).

## 4. Discussion

This study samples the patterns of ABR in bacteria associated with UTIs in symptomatic patients in East Africa. Our main finding is that ABR of the main uropathogens isolated from UTIs (*E. coli*, *Staphylococcus* spp., *Klebsiella* spp., and *Enterococcus* spp.) are severely high. Further, approximately half of the bacterial pathogens isolated from UTIs have MDR. That rate was much higher among inpatients (which we assume are predominantly hospital-acquired UTI) than in outpatients (which we assume are predominantly community-acquired UTIs), as has been described previously [20,21]. These alarming data provide further empirical evidence to enrich the findings of recent studies describing the high morbidity and mortality burden from ABR in Eastern sub-Saharan Africa [2].

The high proportion of MDR in UTI could suggest a previous record of inappropriate AB use in Kenya, Tanzania, and Uganda, which is often considered to be one of the key drivers of AMR. This could be caused by (a) the scarcity of microbiology and AB susceptibility data in this region [2,3,4] which can hamper the management of more appropriate empirical treatment for UTIs, and (b) AB self-treatment and the prevalence of over-the-counter sales of ABs in the community, widespread in LMICs [22,23,24]. Suboptimal management of treatment and the community transmission of MDR bacteria promoted by crowded and less sanitary living conditions, more common in low-and middle-income countries (LMICs) [25], could explain the high proportions of MDR bacteria and the tendency in the study cohort to come straight to clinic [26].

In addition, we found differences among countries, with Kenya presenting lower percentage of MDR bacteria (36.9%) than Tanzania (60.9%) and Uganda (57.5%). Worthy of special attention are the high MDR rates of *E. coli* (>66.0%) found in Tanzania and Uganda, as well as MDR *Klebsiella* (62.2%), *Staphylococcus* (72.3%) and *Enterococcus* (46.6%) species observed in Tanzania, which were much higher than in the other countries. These results emphasise the importance of implementing or reviewing country-specific empirical AB recommendations, which could increase AB efficacy and reduce the burden of AMR according to the resistance rates of each country [27].

Globally, our results fill a crucial data gap which we hope will: (a) feed into guidelines for UTI empirical treatment, (b) provide vital surveillance data for East Africa and indeed the wider Sub-Saharan region, a region with one of the highest ABR-mortality burdens in the world, and c) contribute to development of interventions to monitor and counter the threat of ABR across the region through improved diagnostics and surveillance.

The sparsity of data about the prevalence of resistance for key pathogen–antibiotic combinations in LMICs is a limiting factor for drafting empirical treatment guidelines which can promote appropriate prescription hence hindering the selection of the resistant pathogens [2,3,4]. In this study, we have found a high prevalence of the most insidious AB-pathogen combinations i.e. third-generation cephalosporin (3GC)-resistant *E. coli* (29.3%), fluoroquinolone-resistant *E. coli* (45.8%), 3GC-resistant *Klebsiella* spp. (53.9%), methicillin-resistant staphylococci (39.7%), fluoroquinolone-resistant *Enterococcus* spp. (40.1%), and vancomycin-resistant *Enterococcus* spp. (37.2%). However, we observed systematic variations across country settings, with the Kenyan samples showing the lowest rate of resistance to these ABs, which suggest that recommendations for using a specific empirical AB should be tailored according to each country [27]. The high proportion of fluoroquinolone-resistant *E. coli* and fluoroquinolone-resistant *Enterococcus* spp. found in this study, which are in the top six of the most lethal AB-combination in UTI [7], advise against the empirical use of this AB, whose use in treatment of uncomplicated UTI is no longer recommended by WHO [28,29]. The clinical guidelines of Tanzania and Uganda recommended ciprofloxacin as first or second line ABs for the treatment of uncomplicated UTI in outpatients [30,31,32], which could explain the higher fluoroquinolone resistance observed in these two countries than those observed in Kenya.

Methicillin-resistant *S. aureus* was the most lethal drug-pathogen combination in 2019 in the world [2], being in the top ten of resistance-attributable deaths in UTI [7]. Although in our study staphylococci were not analysed to species level, we found an overall rate of methicillin (cefoxitin) resistance of 39.7%. This contrasts with global estimations in sub-Saharan Africa, which has been recently described as one of the lowest in the world (5%) [2]. Our study has revealed *Staphylococcus* spp. as the second most frequent genus in UTI, which is in line with current evidence that point towards a major role of this species as a common cause of UTI [33,34,35]. Although we cannot rule out contamination with *Staphylococcus* spp. in UTI samples, the fact that nearly 2 of every 3 isolates were MDR, and ≈40% were resistant to cefoxitin, should be considered for managing *Staphylococcus* spp. as true causing agents of UTI.

Amoxicillin/clavulanic acid is among the ABs commonly used to treat uncomplicated UTI. In this study, we found a high level of resistance (37.3%-47.1%) to amoxicillin/clavulanic acid in *Enterobacterales*, which could endanger its future empirical use for treatment of UTIs [36] as happened with amoxicillin alone, whose use in uncomplicated UTI is no longer recommended [29]. In addition, the overall resistance to the folate pathway inhibitor trimethoprim was exceptionally high (53.9%-74.4%) in isolates from order *Enterobacterales*, while resistance to nitrofurantoin was low. This trend has been reported in UTIs worldwide, which has led to the prioritization of the use of nitrofurantoin over trimethoprim as the first-line treatment for UTI [37], including East Africa [30,31,32]. In 2021, however, the WHO added single-agent trimethoprim as a recommendation for the treatment of uncomplicated UTI [29], whose empirical use in East Africa (with a trimethoprim-resistance *E. coli* rate of until 84.1% in Tanzania), would make that AB poorly effective for the treatment of UTI in that region.

The study has some limitations. In the design of the HATUA we endeavoured to provide a consistent study framework across the three countries and the three sites within each country where patients were recruited and their samples were processed and analysed. Standardization of methods and operating procedures were applied across the consortium [5] and used by the Kenyan, Tanzanian, and Ugandan chapters of HATUA. However, even with these in place we cannot rule out that some biases in sampling practices or patient populations studied will have occurred.

Within each country three sites were chosen that had three distinct socio-demographical characteristics and represented a different type of site. This was done in order capture the burden of AMR in UTIs across different community settings in each country. Whilst each country selected sites that were representative of each site type, and provided some level of sociodemographic comparability across countries for the study, there is variation that a study of this scale introduces that means that the populations are not equivalent due to geographic, climatic, ethnic and cultural factors. In this regard we note that across the three countries there are differences in the demographic profiles of the patients recruited. For example, in Kenya more recruitment occurred at higher level health facilities, and the cohort had a greater proportion of patients under the age of 35 years in comparison to those of the other countries. We cannot therefore exclude the introduction of bias that may influence some of the observed microbiological results and some of the differences seen between countries. Recognising this, the interpretation of the results should reflect that they do not necessarily represent true country-level differences across the region, as the sampling within the countries is limited to three sites and is not representative of the countries as a whole.

With such a large, multi-site study, and need for comparability, there have been some inevitable trade-offs between depth and breadth, and as a result for most of the isolates, only their identification to genus level is shown. As samples from outpatients were self-collected, there was a risk of contamination in the samples, which could help explain the high levels of *Staphylococcus* spp. found in this study. Although a wide range of the most commonly used/relevant ABs for UTI in the region was tested, this did not include all possible ABs, which could have led to an underestimation of the true MDR proportions, and therefore our estimates of the burden of AMR on patients with UTIs are conservative.

## 5. Conclusions

This multi-site standardised study describes how approximately half of UTI patients that attended to our recruitment centres in Kenya, Tanzania, and Uganda exhibit MDR bacteria. Several of the most hazardous pathogen-antibiotic combinations (third-generation cephalosporins and fluoroquinolones resistant *E. coli*; methicillin-resistant staphylococci (MRS); third-generation cephalosporin-resistant *K. pneumoniae*; vancomycin-resistant enterococci; multi-drug resistant bacteria) were detected at high proportions in UTI, which severely limits the effectiveness of currently used ABs to treat this common infection. These findings should feed directly into guidelines for empiric AB treatment of UTI in East Africa. More broadly, we emphasize the need for urgent investment in routine AMR surveillance programs, expansion of diagnostic laboratory capacities and diagnostic algorithms to facilitate antimicrobial stewardship and call for greater commitment from policymakers to counter the threat of AMR.

## Data Availability

All data produced in the present work are contained in the manuscript

## HATUA consortium includes everyone listed above, plus

Blandina T. Mmbaga (Kilimanjaro Clinical Research Institute and Kilimanjaro Christian Medical University College, Tanzania), John Mwaniki (KEMRI, Kenya), Stella Neema (Makerere University, Uganda), Joseph R. Mwanga (Catholic University of Health and Allied Sciences, Tanzania), Arun Gonzales Decano (University of St Andrews, UK), V. Anne Smith (University of St Andrews, UK), Alison Elliott (London School of Hygiene & Tropical Medicine, UK), Gibson Kibiki (Africa Research Excellence Fund), David Aanensen (University of Oxford, UK).

## Author contributions

AM: Formal analysis, Data curation, Writing – original draft preparation, Writing – review and editing

SEM: Conceptualisation, Formal analysis, Data curation, Writing – original draft preparation, Writing – review and editing.

XK: Formal analysis, data curation; Writing – review and editing

KK: Conceptualisation, Formal analysis, data curation; Writing – review and editing, supervision

SG: Methodology; Validation; Writing – review and editing

JS: Methodology; Validation; Writing – review and editing

JM: Methodology; Investigation; Data curation; Writing – review and editing

JB: Methodology; Investigation; Data curation

IM: Methodology; Investigation; Data curation

MM: Methodology; Investigation; Data curation

DLG: Data curation

MK: Conceptualisation; Methodology; Writing – review and editing

AGL: Conceptualisation; Methodology; Writing – review and editing

WS: Conceptualisation; Methodology; Writing – review and editing

DJS: Conceptualisation; Methodology; Writing – review and editing

AS: Coordinated the study; Writing – review and editing

JK: Conceptualisation; Methodology; Led data collection in Kenya

BA: Conceptualisation; Methodology; Led data collection in Uganda

MH: Led project conceptualisation; Methodology, Funding acquisition; Writing – review and editing

## Acknowledgements

We thank all the patients who participated in the HATUA study and the field and laboratory teams in each of the three countries.

## Ethical approval

The studies involving human participants were reviewed and approved by University of St Andrews, UK (number MD14548, 10/09/19); National Institute for Medical Research, Tanzania (number 2831, updated 26/07/19); CUHAS/BMC Research Ethics and Review Committee (number CREC /266/2018, updated on 02/2019); Mbeya Medical Research and Ethics Committee (number SZEC- 2439/R.A/V.1/303030); Kilimanjaro Christian Medical College, Tanzania (number 2293, updated 14/08/19); Uganda National Council for Science and Technology (number HS2406, 18/06/18); Makerere University, Uganda (number 514, 25/04/18); and Kenya Medical Research Institute (04/06/19, Scientific and Ethics Review Committee (SERU) number KEMRI/SERU/CMR/P00112/3865 V.1.2). The patients/participants provided their written informed consent to participate in this study.

## Appendix. Supplementary materials

### Method S1. Patient selection/Case definition

The study included adults and children (≥ 2 years) with signs and symptoms of urinary tract infections. Patients attended to the health facilities to seek treatment for UTI-like symptoms or for other causes that gave the doctor reason to believe that they might also (or actually) have a UTI. Specifically, the study enrolled: (a) pregnant women with fever and at least one of the following symptoms, lower abdominal pain, flank/back pain, or strong-smelling urine; (b) pregnant women with at least one urinary symptom, i.e., dysuria, pyuria or haematuria; (c) non-pregnant women or men with fever and at least one of the following symptoms, lower abdominal pain, flank/back pain or strong-smelling urine; (d) non-pregnant women or men with at least one urinary symptom, i.e., frequency, dysuria, pyuria, haematuria or urgency [1,2,3]. In addition, the study enrolled all children aged two years and above that complied with at least one of the following criteria: (a) fever and at least one of: vomiting, abdominal pain, flank/back pain, strong-smelling urine or enuresis; (b) at least one of these urinary symptoms: dysuria, pyuria, hematuria, urgency or frequency; (c) or child with at least two of: costovertebral angle tenderness, abdominal or suprapubic tenderness to palpation, palpable bladder or dribbling/poor stream [4].

### Method S2. Antimicrobial susceptibility testing (AST)

The susceptibility or non-susceptibility (resistance) to the tested antibiotics was determined by using the breakpoints (zone diameter interpretive criteria) indicated in the CLSI guidelines [5] with the following modifications. For nitrofurantoin and linezolid, the European Committee on Antimicrobial Susceptibility Testing [6] breakpoints were used. For *Acinetobacter* spp., susceptibility to amoxicillin/clavulanic acid and trimethoprim was determined using the breakpoints for *Enterobacterales*. For *Staphylococcus* spp., methicillin resistance (non-susceptibility to cefoxitin) was calculated using the breakpoints for *S. epidermidis* and *Staphylococcus* spp. For *Pseudomonas* spp., breakpoints for *Pseudomonas aeruginosa* were applied.

### Method S3. Definition and analysis of Multidrug resistance (MDR)

MDR bacteria were defined as isolates resistant to at least one agent in three or more classes of antimicrobial agents, following the European Centre for Disease Prevention and Control (ECDC) guidelines [7], with some modifications. Thus, nitrofurantoin and trimethoprim, two antibiotics routinely used for treating UTIs that are not included in the ECDC tables [7], were also considered for estimating MDR (Table S2). In addition, for those species/genera not incorporated in the ECDC, i.e. *Salmonella*, *Shigella* and *Streptococcus*, the MDR rates were calculated following the same rule as described above, but considering the resistance to a selected pool of tested antibiotics (Table S2). Those *Staphylococcus* spp. isolates that were resistant to cefoxitin (methicillin-resistant), were not automatically considered MDR, and were subjected to the rule of being resistant to at least one agent in three or more classes of antimicrobial agents. In those isolates with intrinsic resistance to a given antibiotic, such antibiotic was not considered for calculating MDR. By means of this approach, isolates from UTIs were classified as MDR or non-MDR. MDR rates were calculated by considering the number of MDR isolates divided by the total isolates where valid MDR data were obtained.

---

**Table S1.** Patient recruitment sites and healthcare facilities in Kenya, Tanzania, and Uganda.

| Country/site | Number of facilities | Source of funding | Levels recruited from<sup>1</sup> |
| --- | --- | --- | --- |
| **Kenya** |  |  |  |
| Makueni | 1 | Public | 5 |
| Nairobi | 4 | Public and private | 3, 4, 5, 6 |
| Nanyuki | 1 | Public | 4 |
| **Tanzania** |  |  |  |
| Kilimanjaro/Moshi | 3 | Public and private | 2, 3, 5 |
| Mbeya | 2 | Public and private | 3, 4 |
| Mwanza | 5 | Public and private | 2, 3, 5 |
| **Uganda** |  |  |  |
| Mbarara | 3 | Public | 3, 5 |
| Nakapiripirit | 3 | Public | 2, 3 |
| Nakasongola | 3 | Public and private | 3, 4 |

<sup>1</sup> Levels of facilities are identified in each country following the Kenya Health Policy 2014-2013 (http://publications.universalhealth2030.org/uploads/kenya_health_policy_2014_to_2030.pdf), Tanzania fifth health sector strategic plan (HSSP V) 2021-2026 (https://p4h.world/en/node/11813#:~:text=2026%20%7C%20P4H%20Network-,Tanzania%20fifth%20health%20sector%20strategic%20plan%20(HSSPV)%2D%202021%2D2026,coverage%20(UHC)%20by%202030) and the Ugandan Hospital and Health Centre IV census survey 2014
In all three countries, lower levels (1-3) refer to primary care, dispensaries or community health centres. Level 4 typically refers to primary referral facilities or specialist health care facilities. Level 5 (and 6 in Kenyan) are higher level/tertiary facilities.

**Table S2.** Antibiotics considered for MDR calculations.

| **Gram negative** | **Amoxicillin/**<br>**clavulanate**<br>**(AMC)** | **Ampicillin**<br>**(AMP)** | **Ceftazidime/**<br>**Ceftriaxone**<br>**(CAZ/CRO)** | **Ciprofloxacin**<br>**(CIP)** | **Gentamicin**<br>**(GEN)** | **Nitrofurantoin**<br>**(NIT)** | **Trimethoprim**<br>**(TMP)** |
| --- | --- | --- | --- | --- | --- | --- | --- |
| *E. coli* | AMC | AMP | CAZ/CRO | CIP | GEN | NIT | TMP |
| *Shigella spp.* | AMC | AMP | CAZ/CRO | CIP | GEN | NIT | TMP |
| *Proteus spp.* | AMC | AMP | CAZ/CRO | CIP | GEN | NIT | TMP |
| *Salmonella spp.* | AMC | AMP | CAZ/CRO | CIP | GEN | NIT | TMP |
| *Serratia spp.* | - | - | CAZ/CRO | CIP | GEN | NIT | TMP |
| *Klebsiella spp.* | AMC | - | CAZ/CRO | CIP | GEN | NIT | TMP |
| *Citrobacter spp.* | - | - | CAZ/CRO | CIP | GEN | NIT | TMP |
| *Enterobacter spp.* | - | - | CAZ/CRO | CIP | GEN | NIT | TMP |
| *Morganella spp.* | - | - | CAZ/CRO | CIP | GEN | NIT | TMP |
| *Pantoea spp.* | - | - | CAZ/CRO | CIP | GEN | NIT | TMP |
| *Providencia spp.* | - | - | CAZ/CRO | CIP | GEN | NIT | TMP |
| *Acinetobacter spp.* | - | - | CAZ/CRO | CIP | GEN | - | TMP |
| *Pseudomonas spp.* | - | - | CAZ | CIP | GEN | - | - |

| **Gram positive** | **Cefoxitin**<br>**(FOX)** | **Erythromycin**<br>**(ERY)** | **Linezolid**<br>**(LNZ)** | **Ciprofloxacin**<br>**(CIP)** | **Gentamicin**<br>**(GEN)** | **Nitrofurantoin**<br>**(NIT)** | **Trimethoprim**<br>**(TMP)** | **Tetracycline**<br>**(TCY)** | **Vancomycin**<br>**(VAN)** |
| --- | --- | --- | --- | --- | --- | --- | --- | --- | --- |
| *Staphylococcus spp.* | FOX | ERY | - | CIP | GEN | NIT | TMP | TCY | - |
| *Enterococcus spp.* | - | ERY | LNZ | CIP | - | NIT | - | TCY | VAN |
| *Streptococcus spp.* | - | ERY | LNZ | - | - | NIT | - | TCY | VAN |

**Table S3.** Prevalence of UTI according to patient type, gender, hospital level age, and country.

| Variables/Country | N/% with UTI |  |  |  |
| --- | --- | --- | --- | --- |
|  | Kenya<br>n (%)<sup>1</sup> | Tanzania<br>n (%)<sup>1</sup> | Uganda<br>n (%)<sup>1</sup> | All 3 countries<br>n (%)<sup>1</sup> |
| **Patient type** |  |  |  |  |
| Outpatient | 943(53.8) | 974(27.4) | 499(27.5) | 2416(33.9) |
| Inpatient | 84(58.3) | 150(50.0) | 3(33.3) | 237(52.3) |
| $\chi^2$, p value<sup>2</sup> | 0.94, p=0.690 | 67.2, p<0.001 | 0.1, p=1.000 | 62.48, p<0.001 |
| **Gender** |  |  |  |  |
| Male | 111(31.9) | 305(27.8) | 64(21.7) | 480(27.6) |
| Female | 916(59.1) | 818(29.7) | 438(28.6) | 2172(37.2) |
| $\chi^2$, p value<sup>2</sup> | 69.1, p<0.001 | 15.2, p=0.004 | 7.7, p=0.217 | 190.6, p<0.001 |
| **Hospital level** |  |  |  |  |
| Level 2 | 0 | 88(28.5) | 126(32.0) | 214(30.4) |
| Level 3 | 162(42.2) | 585(27.2) | 259(25.3) | 1006(28.3) |
| Level 4 | 336(69.1) | 104(28.4) | 37(25.9) | 477(47.9) |
| Level 5/6 | 529(51.4) | 347(33.9) | 80(30.4) | 956(41.3) |
| $\chi^2$, p value<sup>2</sup> | 83.6, p<0.001 | 3.8, p=0.312 | 5.6, p=0.14 | 56.7, p<0.001 |
| **Age** |  |  |  |  |
| <18 | 51(64.6) | 75(21.9) | 11(17.7) | 137(28.3) |
| 18-24 | 286(58.0) | 211(29.4) | 155(27.1) | 652(36.5) |
| 25-34 | 436(52.1) | 243(25.6) | 159(27.8) | 838(35.5) |
| 35-44 | 148(50.3) | 150(27.3) | 75(24.7) | 373(32.5) |
| 45-54 | 49(50.5) | 111(26.1) | 62(35.8) | 222(31.9) |
| 55-64 | 19(44.2) | 95(29.6) | 17(23.9) | 131(30.1) |
| 65-74 | 23(65.7) | 111(39.5) | 12(27.9) | 146(40.7) |
| 75 and above | 15(75.0) | 127(48.7) | 9(40.9) | 151(49.8) |
| $\chi^2$, p value<sup>2</sup> | 17.2, p=0.045 | 80.2, p<0.001 | 12.7, p=0.22 | 56.6, p<0.001 |
| **Total** | 1027(54.1) | 1124(29.2) | 502(27.5) | 2653(35.0) |

<sup>1</sup> % is the prevalence of UTI positive samples, calculated by dividing the number of UTI positive samples by the total number of urine samples cultured (n). A sample was considered UTI-positive if presented $>10^4$ CFU/mL of one or two uropathogens.
<sup>2</sup> Ch-squared testing adjusted for false discovery rate (as described in the methods).

**Table S4.** Distribution of significant microorganisms isolated from specimens of symptomatic patients with UTI (UTI positive patients), according to the country and the type of patient (Outpatient or Inpatient).

| Country | Kenya Outpatient |  |  | Kenya Inpatient |  |  | Tanzania Outpatient |  |  | Tanzania Inpatient |  |  | Uganda Outpatient |  |  | Uganda Inpatient |  |  | All 3 countries Outpatient |  |  | All 3 countries Inpatient |  |  |
| --- | --- | --- | --- | --- | --- | --- | --- | --- | --- | --- | --- | --- | --- | --- | --- | --- | --- | --- | --- | --- | --- | --- | --- | --- |
| Microbial Isolates | n | %<sup>1</sup> | Prev (%)<sup>2</sup> | n | % | Prev (%) | n | % | Prev (%) | n | % | Prev (%) | n | % | Prev (%) | n | % | Prev (%) | n | % | Prev (%) | n | % | Prev (%) |
| *Escherichia coli* | 294 | 31.2 | 16.9 | 23 | 27.4 | 16.0 | 350 | 35.9 | 9.9 | 52 | 34.7 | 17.3 | 262 | 52.5 | 14.4 | 1 | 33.3 | 11.1 | 906 | 37.5 | 12.8 | 76 | 32.1 | 16.8 |
| *Klebsiella* spp. | 0.0 | 0.0 | 0.0 | 0.0 | 0.0 | 0.0 | 64 | 6.6 | 1.8 | 26 | 17.3 | 8.7 | 63 | 12.6 | 3.5 | 1 | 33.3 | 11.1 | 127 | 5.3 | 1.8 | 27 | 11.4 | 6.0 |
| *Proteus* spp. | 66 | 7.0 | 3.8 | 3 | 3.6 | 2.1 | 12 | 1.2 | 0.3 | 1 | 0.7 | 0.3 | 15 | 3.0 | 0.8 | 0 | 0.0 | 0.0 | 93 | 3.8 | 1.3 | 4 | 1.7 | 0.9 |
| *Acinetobacter* spp. | 7 | 0.7 | 0.4 | 1 | 1.2 | 0.7 | 14 | 1.4 | 0.4 | 5 | 3.3 | 1.7 | 5 | 1.0 | 0.3 | 0 | 0.0 | 0.0 | 26 | 1.1 | 0.4 | 6 | 2.5 | 1.3 |
| *Pseudomonas aeruginosa* | 6 | 0.6 | 0.3 | 2 | 2.4 | 1.4 | 25 | 2.6 | 0.7 | 16 | 10.7 | 5.3 | 1 | 0.2 | 0.1 | 0 | 0.0 | 0.0 | 32 | 1.3 | 0.4 | 18 | 7.6 | 4.0 |
| Miscellaneous Gram-negative<sup>3</sup> | 87 | 9.2 | 5.0 | 14 | 16.7 | 9.7 | 94 | 9.7 | 2.6 | 17 | 11.3 | 5.7 | 51 | 10.2 | 2.8 | 1 | 33.3 | 11.1 | 232 | 9.6 | 3.3 | 32 | 13.5 | 7.1 |
| *Staphylococcus* spp. | 359 | 38.1 | 20.6 | 28 | 33.3 | 19.4 | 205 | 21.0 | 5.8 | 15 | 10.0 | 5.0 | 91 | 18.2 | 5.0 | 0 | 0.0 | 0.0 | 655 | 27.1 | 9.2 | 43 | 18.1 | 9.5 |
| *Enterococcus* spp. | 81 | 8.6 | 4.7 | 5 | 6.0 | 3.5 | 47 | 4.8 | 1.3 | 11 | 7.3 | 3.7 | 3 | 0.6 | 0.2 | 0 | 0.0 | 0.0 | 131 | 5.4 | 1.9 | 16 | 6.8 | 3.5 |
| Miscellaneous Gram-positive<sup>4</sup> | 28 | 3.0 | 1.6 | 3 | 3.6 | 2.1 | 54 | 5.5 | 1.5 | 1 | 0.7 | 0.3 | 8 | 1.6 | 0.4 | 0 | 0.0 | 0.0 | 90 | 3.7 | 1.3 | 4 | 1.7 | 0.9 |
| Yeast | 2 | 0.2 | 0.1 | 0 | 0.0 | 0.0 | 25 | 2.6 | 0.7 | 3 | 2.0 | 1.0 | 0 | 0.0 | 0.0 | 0 | 0.0 | 0.0 | 27 | 1.1 | 0.4 | 3 | 1.3 | 0.7 |
| Missing species data | 13 | 1.4 | 0.7 | 5 | 6.0 | 3.5 | 84 | 8.6 | 2.4 | 3 | 2.0 | 1.0 | 0 | 0.0 | 0.0 | 0 | 0.0 | 0.0 | 97 | 4.0 | 1.4 | 8 | 3.4 | 1.8 |
| Total | 943 | 100 | 53.8 | 84 | 100 | 58.3 | 974 | 100 | 27.4 | 150 | 100 | 50.0 | 499 | 100 | 27.5 | 3 | 100 | 33.3 | 2416 | 100 | 33.9 | 237 | 100 | 52.3 |

<sup>1</sup>% = percentage of isolates corresponding to that species, from that country and patient type (for example in the first column, calculated by 294/943\*100)
<sup>2</sup>Prev = prevalence proportion (e.g. number of *E. coli* isolates with respect to the total number of urine specimens that were cultured in that country, for that type of patient. For example, the third column is calculated by 294/1752\*100.
<sup>3</sup>This comprises *Aeromonas* spp. (n=1), *Citrobacter* spp. (n=16), *Enterobacter* spp. (n=24), *Moraxella* spp. (n=1), *Morganella* spp. (n=6), *Pantoea* spp. (n=2), *Providencia* spp. (n=2), *Salmonella* spp. (n=2), *Serratia* spp. (n=4), *Shigella* spp. (n=1), *Stenotrophomonas* spp. (n=1), and undetermined Gram-negative bacteria (n=115)
<sup>4</sup>This comprises *Bacillus* spp. (n=19), *Clostridium* spp. (n=1), *Corynebacterium* spp. (n=1), *Lactobacillus* spp. (n=6), *Streptococcus* spp. (n=50), and undetermined Gram-positive bacteria (n=16).

**Table S5.** Antibiotic susceptibility, ESBL and MDR rates of *Enterobacterales*

|  | *Escherichia coli* |  |  |  |  |  |  |  | *Klebsiella* spp. |  |  |  |  |  |  |  | Other *Enterobacterales*<sup>1</sup> |  |  |  |  |  |  |  |
| --- | --- | --- | --- | --- | --- | --- | --- | --- | --- | --- | --- | --- | --- | --- | --- | --- | --- | --- | --- | --- | --- | --- | --- | --- |
|  | Kenya |  | Tanzania |  | Uganda |  | Three countries |  | Kenya |  | Tanzania |  | Uganda |  | Three countries |  | Kenya |  | Tanzania |  | Uganda |  | Three countries |  |
|  | %<sup>2</sup> | n<sup>3</sup> | % | n | % | n | % | n | % | n | % | n | % | n | % | n | % | n | % | n | % | n | % | n |
| **Ampicillin** | 21.8 | 317 | 87.1 | 402 | 81.7 | 262 | 64.5 | 981 | NA | NA | 92.2 | 90 | 95.3 | 64 | 93.5 | 154 | 11.5 | 78 | 84.3 | 51 | 80.0 | 25 | 46.8 | 154 |
| **Amoxicillin-clavulanic acid** | 17.4 | 316 | 43.0 | 402 | 56.3 | 252 | 38.1 | 970 | NA | NA | 58.9 | 90 | 61.9 | 63 | 60.1 | 153 | 10.3 | 78 | 64.7 | 51 | 66.7 | 24 | 37.3 | 153 |
| **Ceftazidime** | 8.5 | 317 | 28.1 | 402 | 31.7 | 252 | 22.7 | 971 | NA | NA | 47.8 | 90 | 31.7 | 63 | 41.2 | 153 | 3.8 | 78 | 23.5 | 51 | 37.5 | 24 | 15.7 | 153 |
| **Ceftriaxone** | 9.2 | 315 | 32.1 | 402 | 46.2 | 262 | 28.5 | 979 | NA | NA | 52.2 | 90 | 50.0 | 64 | 51.3 | 154 | 5.1 | 78 | 25.5 | 51 | 56.0 | 25 | 20.1 | 154 |
| **Ciprofloxacin** | 30.0 | 317 | 57.0 | 402 | 47.9 | 263 | 45.8 | 982 | NA | NA | 55.6 | 90 | 26.6 | 64 | 43.5 | 154 | 33.8 | 77 | 45.1 | 51 | 44.0 | 25 | 39.2 | 153 |
| **Gentamicin** | 16.9 | 314 | 27.6 | 402 | 19.0 | 184 | 22.1 | 900 | NA | NA | 31.1 | 90 | 10.3 | 39 | 24.8 | 129 | 11.8 | 76 | 21.6 | 51 | 7.1 | 14 | 14.9 | 141 |
| **Nitrofurantoin** | 2.9 | 313 | 6.7 | 402 | 1.5 | 263 | 4.1 | 978 | NA | NA | 18.9 | 90 | 7.8 | 64 | 14.3 | 154 | 10.3 | 78 | 31.4 | 51 | 44.0 | 25 | 22.7 | 154 |
| **Trimethoprim** | 57.9 | 316 | 84.1 | 402 | 79.5 | 263 | 74.4 | 981 | NA | NA | 68.9 | 90 | 54.7 | 64 | 63.0 | 154 | 65.4 | 78 | 62.7 | 51 | 52.0 | 25 | 62.3 | 154 |
| **ESBL<sup>4</sup>** | 9.5 | 317 | 32.6 | 402 | 48.3 | 263 | 29.3 | 982 | NA | NA | 53.3 | 90 | 54.7 | 64 | 53.9 | 154 | 6.4 | 78 | 29.4 | 51 | 56.0 | 25 | 22.1 | 154 |
| **MDR<sup>5</sup>** | 22.7 | 317 | 66.4 | 402 | 66.2 | 263 | 52.2 | 982 | NA | NA | 62.2 | 90 | 34.4 | 64 | 50.6 | 154 | 19.2 | 78 | 43.1 | 51 | 44.0 | 25 | 31.2 | 154 |

<sup>1</sup> Other *Enterobacterales* includes *Citrobacter*, *Enterobacter*, *Morganella*, *Proteus*, *Providencia*, *Pantoea*, *Salmonella*, *Serratia* and *Shigella* species.
<sup>2</sup> % is the frequency of non-susceptible isolates, expressed in percentage.
<sup>3</sup> n is the total number of isolates that were tested for a specific antibiotic.
<sup>4</sup> Possible producers of extended-spectrum β-lactamase (ESBL) were determined by considering the resistance to antibiotics Ceftazidime and Ceftriaxone, according to CLSI guidelines [17].
<sup>5</sup> Multi drug-resistance (MDR) was defined as non-susceptibility to at least one antimicrobial agent in three or more antimicrobial categories, according to the ECDC guidelines [10] with some modifications, as described in the methods section.

**Table S6.** Antibiotic susceptibility, ESBL and MDR rates of relevant *Enterobacterales* isolated from outpatients with UTI.

|  | *Escherichia coli* (outpatient) |  |  |  |  |  |  |  | *Klebsiella* spp. (outpatient) |  |  |  |  |  |  |  | Other *Enterobacterales*<sup>1</sup> (outpatient) |  |  |  |  |  |  |  |
| --- | --- | --- | --- | --- | --- | --- | --- | --- | --- | --- | --- | --- | --- | --- | --- | --- | --- | --- | --- | --- | --- | --- | --- | --- |
| Country | Kenya |  | Tanzania |  | Uganda |  | Three countries |  | Kenya |  | Tanzania |  | Uganda |  | Three countries |  | Kenya |  | Tanzania |  | Uganda |  | Three countries |  |
|  | %<sup>2</sup> | n<sup>3</sup> | % | n | % | n | % | n | % | n | % | n | % | n | % | n | % | n | % | n | % | n | % | n |
| **Ampicillin** | 21.1 | 294 | 85.7 | 350 | 81.6 | 261 | 63.5 | 905 | NA | NA | 93.8 | 64 | 95.2 | 63 | 94.5 | 127 | 11.0 | 73 | 82.5 | 40 | 80 | 25 | 44.2 | 138 |
| **Amoxicillin/**<br>**clavulanic acid** | 17.1 | 293 | 39.1 | 350 | 56.6 | 251 | 36.8 | 894 | NA | NA | 54.7 | 64 | 61.3 | 62 | 57.9 | 126 | 9.6 | 73 | 57.5 | 40 | 66.7 | 24 | 33.6 | 137 |
| **Ceftazidime** | 7.5 | 294 | 22.6 | 350 | 31.9 | 251 | 20.2 | 895 | NA | NA | 35.9 | 64 | 30.6 | 62 | 33.3 | 126 | 1.4 | 73 | 15.0 | 40 | 37.5 | 24 | 11.7 | 137 |
| **Ceftriaxone** | 8.2 | 293 | 26.0 | 350 | 46.0 | 261 | 26.0 | 904 | NA | NA | 42.2 | 64 | 49.2 | 63 | 45.7 | 127 | 2.7 | 73 | 17.5 | 40 | 56.0 | 25 | 16.7 | 138 |
| **Ciprofloxacin** | 29.6 | 294 | 54.0 | 350 | 47.7 | 262 | 44.3 | 906 | NA | NA | 50 | 64 | 25.4 | 63 | 37.8 | 127 | 31.9 | 72 | 42.5 | 40 | 44.0 | 25 | 37.2 | 137 |
| **Gentamicin** | 14.7 | 292 | 24.3 | 350 | 19.0 | 184 | 19.7 | 826 | NA | NA | 23.4 | 64 | 10.3 | 39 | 18.4 | 103 | 9.9 | 71 | 15.0 | 40 | 7.1 | 14 | 11.2 | 125 |
| **Nitrofurantoin** | 2.1 | 291 | 4.9 | 350 | 1.5 | 262 | 3.0 | 903 | NA | NA | 15.6 | 64 | 7.9 | 63 | 11.8 | 127 | 11.0 | 73 | 32.5 | 40 | 44.0 | 25 | 23.2 | 138 |
| **Trimethoprim** | 59.0 | 293 | 83.4 | 350 | 79.4 | 262 | 74.4 | 905 | NA | NA | 60.9 | 64 | 54.0 | 63 | 57.5 | 127 | 64.4 | 73 | 62.5 | 40 | 52.0 | 25 | 61.6 | 138 |
| **ESBL<sup>4</sup>** | 8.5 | 294 | 26.6 | 350 | 48.1 | 262 | 26.9 | 906 | NA | NA | 43.8 | 64 | 54.0 | 63 | 48.8 | 127 | 2.7 | 73 | 22.5 | 40 | 56.0 | 25 | 18.1 | 138 |
| **MDR<sup>5</sup>** | 21.4 | 294 | 64.0 | 350 | 66.0 | 262 | 50.8 | 906 | NA | NA | 56.3 | 64 | 33.3 | 63 | 44.9 | 127 | 16.4 | 73 | 40 | 40 | 44.0 | 25 | 28.3 | 138 |

<sup>1</sup> Other *Enterobacterales* includes *Citrobacter*, *Enterobacter*, *Morganella*, *Proteus*, *Providencia*, *Pantoea*, *Salmonella*, *Serratia* and *Shigella* species.
<sup>2</sup> % is the frequency of non-susceptible isolates, expressed in percentage.
<sup>3</sup> n is the total number of isolates that were tested for a specific antibiotic.
<sup>4</sup> Possible producers of extended-spectrum β-lactamase (ESBL) were determined by considering the resistance to antibiotics Ceftazidime and Ceftriaxone, according to CLSI guidelines [17].
<sup>5</sup> Multi drug-resistance (MDR) was defined as non-susceptibility to at least one antimicrobial agent in three or more antimicrobial categories, according to the ECDC guidelines [10].

**Table S7.** Antibiotic susceptibility, ESBL and MDR rates of relevant *Enterobacterales* isolated from inpatients with UTI.

|  | *Escherichia coli* (inpatient) |  |  |  |  |  |  |  | *Klebsiella* spp. (inpatient) |  |  |  |  |  |  |  | Other *Enterobacterales*<sup>1</sup> (inpatient) |  |  |  |  |  |
| --- | --- | --- | --- | --- | --- | --- | --- | --- | --- | --- | --- | --- | --- | --- | --- | --- | --- | --- | --- | --- | --- | --- |
| Country | Kenya |  | Tanzania |  | Uganda |  | Three countries |  | Kenya |  | Tanzania |  | Uganda |  | Three countries |  | Kenya |  | Tanzania |  | Three Countries<sup>2</sup> |  |
|  | %<sup>3</sup> | n<sup>4</sup> | % | n | % | n | % | n | % | n | % | n | % | n | % | n | % | n | % | n | % | n |
| Ampicillin | 30.4 | 23 | 96.2 | 52 | 100 | 1 | 76.3 | 76 | NA | NA | 88.5 | 26 | 100 | 1 | 88.9 | 27 | 20 | 5 | 90.9 | 11 | 68.8 | 16 |
| Amoxicillin/<br>clavulanic acid | 21.7 | 23 | 69.2 | 52 | 0 | 1 | 53.9 | 76 | NA | NA | 69.2 | 26 | 100 | 1 | 70.4 | 27 | 20 | 5 | 90.9 | 11 | 68.8 | 16 |
| Ceftazidime | 21.7 | 23 | 65.4 | 52 | 0 | 1 | 51.3 | 76 | NA | NA | 76.9 | 26 | 100 | 1 | 77.8 | 27 | 40 | 5 | 54.5 | 11 | 50 | 16 |
| Ceftriaxone | 22.7 | 22 | 73.1 | 52 | 100 | 1 | 58.7 | 75 | NA | NA | 76.9 | 26 | 100 | 1 | 77.8 | 27 | 40 | 5 | 54.5 | 11 | 50 | 16 |
| Ciprofloxacin | 34.8 | 23 | 76.9 | 52 | 100 | 1 | 64.5 | 76 | NA | NA | 69.2 | 26 | 100 | 1 | 70.4 | 27 | 60 | 5 | 54.5 | 11 | 56.3 | 16 |
| Gentamicin | 45.5 | 22 | 50 | 52 | NA<sup>5</sup> | NA | 48.6 | 74 | NA | NA | 50 | 26 | NA | NA | 50.0 | 26 | 40 | 5 | 45.5 | 11 | 43.8 | 16 |
| Nitrofurantoin | 13.6 | 22 | 19.2 | 52 | 0 | 1 | 17.3 | 75 | NA | NA | 26.9 | 26 | 0 | 1 | 25.9 | 27 | 0 | 5 | 27.3 | 11 | 18.8 | 16 |
| Trimethoprim | 43.5 | 23 | 88.5 | 52 | 100 | 1 | 75.0 | 76 | NA | NA | 88.5 | 26 | 100 | 1 | 88.9 | 27 | 80 | 5 | 63.6 | 11 | 68.8 | 16 |
| ESBL<sup>6</sup> | 21.7 | 23 | 73.1 | 52 | 100 | 1 | 57.9 | 76 | NA | NA | 76.9 | 26 | 100 | 1 | 77.8 | 27 | 60 | 5 | 54.5 | 11 | 56.3 | 16 |
| MDR<sup>7</sup> | 39.1 | 23 | 82.7 | 52 | 100 | 1 | 69.7 | 76 | NA | NA | 76.9 | 26 | 100 | 1 | 77.8 | 27 | 60 | 5 | 54.5 | 11 | 56.3 | 16 |

<sup>1</sup> Other *Enterobacterales* includes *Citrobacter*, *Enterobacter*, *Morganella*, *Proteus*, *Providencia*, *Pantoea*, *Salmonella*, *Serratia* and *Shigella* species.
<sup>2</sup> No *Enterobacterales* were isolated from Uganda inpatients
<sup>3</sup> % is the frequency of non-susceptible isolates, expressed in percentage.
<sup>4</sup> n is the total number of isolates that were tested for a specific antibiotic.
<sup>5</sup> NA: not applicable.
<sup>6</sup> Possible producers of extended-spectrum β-lactamase (ESBL) were determined by considering the resistance to antibiotics Ceftazidime and Ceftriaxone, according to CLSI guidelines [17].
<sup>7</sup> Multi drug-resistance (MDR) was defined as non-susceptibility to at least one antimicrobial agent in three or more antimicrobial categories, according to the ECDC guidelines [10] with some modifications, as described in the methods section.

**Table S8.**
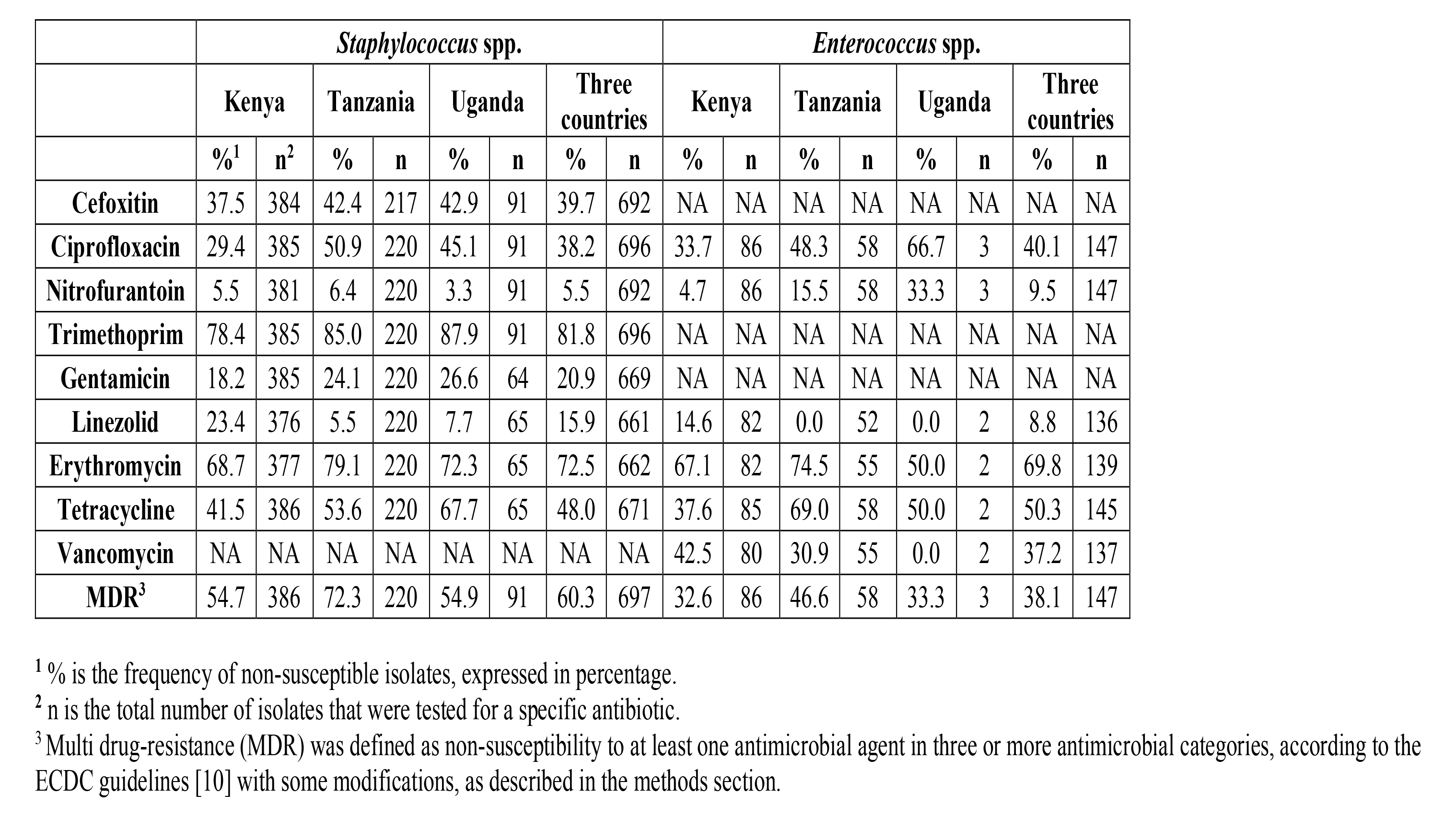
Antibiotic susceptibility and MDR rates of relevant Gram-positive uropathogens.

|  | *Staphylococcus spp.* |  |  |  |  |  |  |  | *Enterococcus spp.* |  |  |  |  |  |  |  |
| --- | --- | --- | --- | --- | --- | --- | --- | --- | --- | --- | --- | --- | --- | --- | --- | --- |
|  | Kenya |  | Tanzania |  | Uganda |  | Three countries |  | Kenya |  | Tanzania |  | Uganda |  | Three countries |  |
|  | %<sup>1</sup> | n<sup>2</sup> | % | n | % | n | % | n | % | n | % | n | % | n | % | n |
| **Cefoxitin** | 37.5 | 384 | 42.4 | 217 | 42.9 | 91 | 39.7 | 692 | NA | NA | NA | NA | NA | NA | NA | NA |
| **Ciprofloxacin** | 29.4 | 385 | 50.9 | 220 | 45.1 | 91 | 38.2 | 696 | 33.7 | 86 | 48.3 | 58 | 66.7 | 3 | 40.1 | 147 |
| **Nitrofurantoin** | 5.5 | 381 | 6.4 | 220 | 3.3 | 91 | 5.5 | 692 | 4.7 | 86 | 15.5 | 58 | 33.3 | 3 | 9.5 | 147 |
| **Trimethoprim** | 78.4 | 385 | 85.0 | 220 | 87.9 | 91 | 81.8 | 696 | NA | NA | NA | NA | NA | NA | NA | NA |
| **Gentamicin** | 18.2 | 385 | 24.1 | 220 | 26.6 | 64 | 20.9 | 669 | NA | NA | NA | NA | NA | NA | NA | NA |
| **Linezolid** | 23.4 | 376 | 5.5 | 220 | 7.7 | 65 | 15.9 | 661 | 14.6 | 82 | 0.0 | 52 | 0.0 | 2 | 8.8 | 136 |
| **Erythromycin** | 68.7 | 377 | 79.1 | 220 | 72.3 | 65 | 72.5 | 662 | 67.1 | 82 | 74.5 | 55 | 50.0 | 2 | 69.8 | 139 |
| **Tetracycline** | 41.5 | 386 | 53.6 | 220 | 67.7 | 65 | 48.0 | 671 | 37.6 | 85 | 69.0 | 58 | 50.0 | 2 | 50.3 | 145 |
| **Vancomycin** | NA | NA | NA | NA | NA | NA | NA | NA | 42.5 | 80 | 30.9 | 55 | 0.0 | 2 | 37.2 | 137 |
| **MDR<sup>3</sup>** | 54.7 | 386 | 72.3 | 220 | 54.9 | 91 | 60.3 | 697 | 32.6 | 86 | 46.6 | 58 | 33.3 | 3 | 38.1 | 147 |

<sup>1</sup> % is the frequency of non-susceptible isolates, expressed in percentage.
<sup>2</sup> n is the total number of isolates that were tested for a specific antibiotic.
<sup>3</sup> Multi drug-resistance (MDR) was defined as non-susceptibility to at least one antimicrobial agent in three or more antimicrobial categories, according to the ECDC guidelines [10] with some modifications, as described in the methods section.

**Table S9.** Antibiotic susceptibility, ESBL and MDR rates of *Staphylococcus* spp. and *Enterococcus* spp. isolated from outpatients with UTI.

|  | *Staphylococcus* spp. (outpatient) |  |  |  |  |  |  |  | *Enterococcus* spp. (outpatient) |  |  |  |  |  |  |  |
| --- | --- | --- | --- | --- | --- | --- | --- | --- | --- | --- | --- | --- | --- | --- | --- | --- |
| Country | Kenya |  | Tanzania |  | Uganda |  | Three countries |  | Kenya |  | Tanzania |  | Uganda |  | Three countries |  |
|  | %<sup>1</sup> | n<sup>2</sup> | % | n | % | n | % | n | % | n | % | n | % | n | % | n |
| **Cefoxitin** | 35.1 | 356 | 43.1 | 202 | 42.9 | 91 | 38.7 | 649 | NA<sup>3</sup> | NA | NA | NA | NA | NA | NA | NA |
| **Ciprofloxacin** | 28.3 | 357 | 50.7 | 205 | 45.1 | 91 | 37.7 | 653 | 32.1 | 81 | 42.6 | 47 | 66.7 | 3 | 36.6 | 131 |
| **Nitrofurantoin** | 4.5 | 353 | 5.9 | 205 | 3.3 | 91 | 4.8 | 649 | 4.9 | 81 | 19.1 | 47 | 33.3 | 3 | 10.7 | 131 |
| **Trimethoprim** | 77.9 | 357 | 85.4 | 205 | 87.9 | 91 | 81.6 | 653 | NA | NA | NA | NA | NA | NA | NA | NA |
| **Gentamicin** | 15.1 | 357 | 23.4 | 205 | 26.6 | 64 | 19.0 | 626 | NA | NA | NA | NA | NA | NA | NA | NA |
| **Linezolid** | 20.9 | 349 | 4.9 | 205 | 7.7 | 65 | 14.2 | 619 | 15.6 | 77 | 0.0 | 41 | 0.0 | 2 | 10.0 | 120 |
| **Erythromycin** | 67.4 | 350 | 78.0 | 205 | 72.3 | 65 | 71.5 | 620 | 64.9 | 77 | 70.5 | 44 | 50.0 | 2 | 66.7 | 123 |
| **Tetracycline** | 40.8 | 358 | 53.2 | 205 | 67.7 | 65 | 47.6 | 628 | 35.0 | 80 | 63.8 | 47 | 50.0 | 2 | 45.7 | 129 |
| **Vancomycin** | NA | NA | NA | NA | NA | NA | NA | NA | 41.3 | 75 | 29.5 | 44 | 0.0 | 2 | 36.4 | 121 |
| **MDR<sup>4</sup>** | 53.1 | 358 | 72.7 | 205 | 54.9 | 91 | 59.5 | 654 | 30.9 | 81 | 40.4 | 47 | 33.3 | 3 | 34.4 | 131 |

<sup>1</sup> % is the frequency of non-susceptible isolates, expressed in percentage.
<sup>2</sup> n is the total number of isolates that were tested for a specific antibiotic.
<sup>3</sup> NA: not applicable.
<sup>4</sup> Multi drug-resistance (MDR) was defined as non-susceptibility to at least one antimicrobial agent in three or more antimicrobial categories, according to the ECDC guidelines [10] with some modifications, as described in the methods section.

**Table S10.**
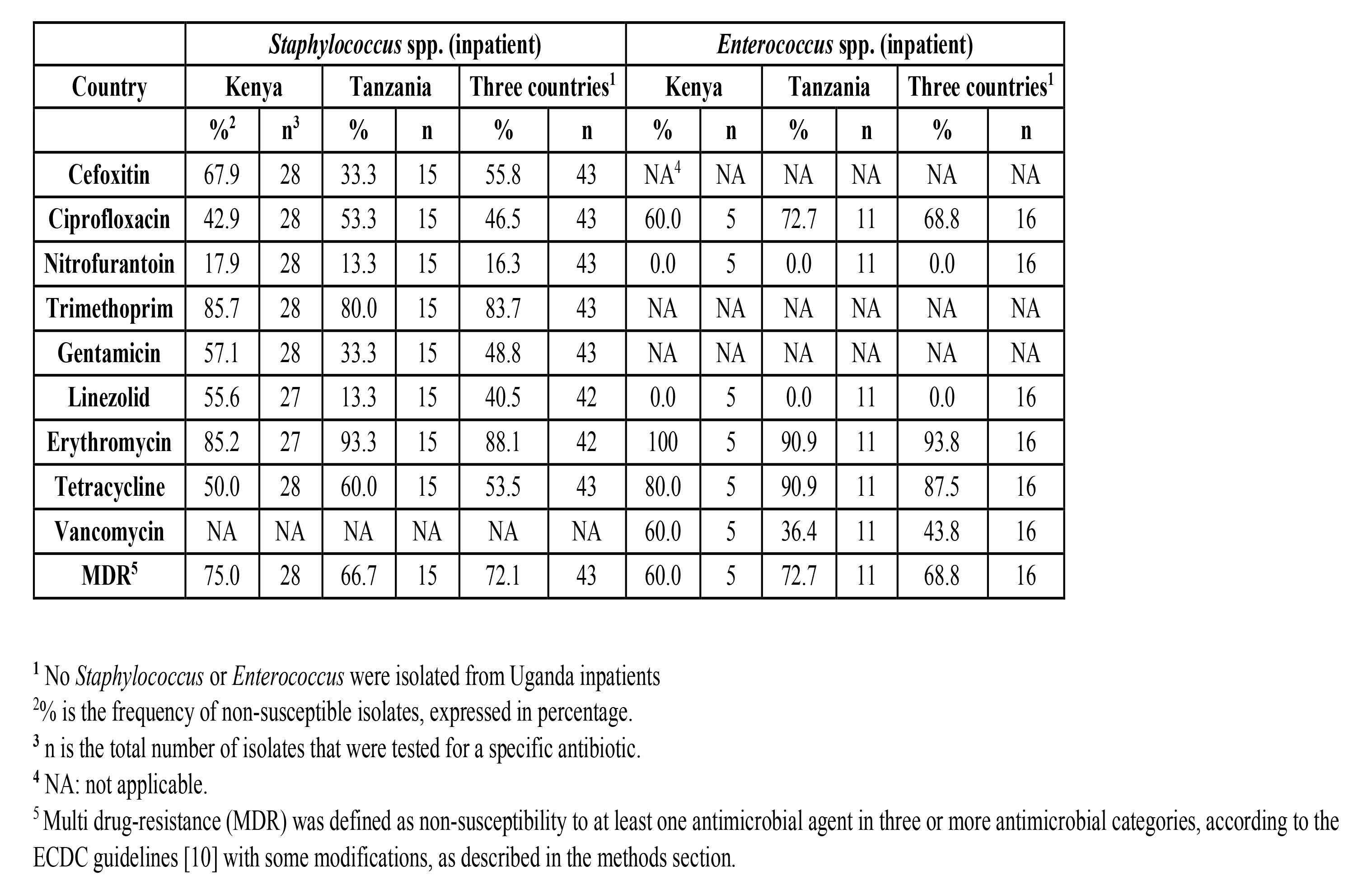
Antibiotic susceptibility, ESBL and MDR rates of *Staphylococcus* spp. and *Enterococcus* spp. isolated from inpatients with UTI.

